# Sources and transmission dynamics of *Campylobacter* and *Shigella* using culture-independent molecular methods in Tanzania (SCAT): a protocol for a prospective observational cohort study

**DOI:** 10.64898/2026.06.23.26356359

**Authors:** Peter M. Mbelele, Sifaeli Katengu, Erin G. Wettstone, Suporn Pholwat, Sarah Elwood, Tahmina Ahmed, Godfrey W. Guga, Mariamu Temu, Fredrick Habiye, Caroline Kimathi, Kelvin Msoka, Restituta Mosha, Laura H Kwong, Andrew F. Brouwer, Joseph N.S. Eisenberg, Elizabeth T. Rogawski McQuade, Mami Taniuchi, Esto Mduma, James A. Platts-Mills

## Abstract

**Introduction:** Enteric infections caused by *Shigella* and *Campylobacter* species are associated with acute diarrhea as well as enteric dysfunction leading to chronic malnutrition and poor child development. However, interventional studies to reduce exposure to these pathogens have been ineffective, and evidence that identifies putative sources and species transmission dynamics between humans, animals and the environment is scarce. Because isolation of these organisms from both human and environmental samples is challenging, studies using molecular detection may help reduce this substantial knowledge gap and lead to targeted interventions to reduce the burden of disease.

**Methods and analysis:** This longitudinal cohort study will enroll 100 index infants as well as their families in Haydom, Tanzania. Families will be followed for one year, with collection of stool samples monthly and during incident diarrhea from all household members.

Additionally, we will sample from the index child’s environment, including domestic animal stools, drinking water, milk, porridge, flies, and soil. All samples will undergo nucleic acid extraction and quantitative PCR for *Campylobacter* and *Shigella*. Serial serologic tests will also be performed to identify seroconversion to these pathogens. Associations between pathogen detection from the environment and household members and detection in index children will be estimated, and the temporal patterning of infection cases will be analyzed using transmission models.

**Ethics and dissemination:** This trial has been approved by the Tanzanian National Institute for Medical Research and the University of Virginia Institutional Review Board.

**STRENGTHS AND LIMITATIONS OF THIS STUDY:**

- This study will apply molecular diagnostics to identify *Shigella* and *Campylobacter* infections in a closely-followed cohort of index children and their families in a high-burden setting while contemporaneously sampling the household environment to identify potential sources of exposure.
- The interrogation of a broad range of potential sources and transmission pathways for both *Shigella* (for which person-to-person transmission is thought to be primary) and *Campylobacter* (for which environment-to-person transmission is thought to be primary) will allow us to test long-held assumptions about risk factors for and interventions to reduce the burden of disease associated with these organisms.
- Molecular diagnostics should increase the yield of pathogen detection from a broad range of environmental sources, but careful analytical consideration will be needed to due to the high sensitivity of this approach.

## INTRODUCTION

Diarrhea is the second leading cause of death in children under five years, accounting for annual mortality of approximately 500,000 globally, with the majority of these deaths occurring in sub-Saharan Africa^[1]^. In addition to acute mortality and morbidity, early and repeated enteric infections have been associated with enteric dysfunction, chronic malnutrition, and impaired cognitive development^[2]^; *Shigella* and *Campylobacter* are the two enteric pathogens that have most consistently associated with these long-term outcomes^[3–5]^. Improvements in water, sanitation, and hygiene are broadly considered to be an important approach to reducing the burden of these pathogens; however, recent large cluster-randomized trials have shown little reduction, particularly of bacterial infections ^[6,7]^.

While the general routes of exposure to enteric pathogens through fecal-oral contact are well known^[8,9]^, there are critical limitations to our understanding of pathogen-specific transmission pathways, especially in young children in low-resource settings where the burden and consequences are highest^[10]^. Isolation of bacterial enteropathogens such as *Campylobacter* and *Shigella* from stool, food, water or other environmental samples using conventional culture media bears significant technical challenges including low sensitivity^[11–13]^. The application of molecular diagnostic detection methods could potentially address these limitations^[14]^.

*Shigella* and *Campylobacter* are model organisms to interrogate transmission ecology because they are thought to lie on opposite ends of the continuum of person-to-person (*Shigella*)^[15]^ to environment-to-person (*Campylobacter*) transmission^[16]^. Estimating the degree to which *Campylobacter* transmission is person-to-person and *Shigella* transmission is environment-to-person will significantly advance our understanding of enteric pathogen transmission dynamics by moving from a broad understanding of fecal-oral transmission routes to pathogen-specific household and environmental pathways of highest relevance. We thus propose to study household and environmental transmission of these two important pathogens by close follow-up of a longitudinal cohort of index children and their families, with frequent collection of human stool, animal feces, and environmental samples, using molecular detection methods to identify *Campylobacter* and *Shigella* in the high-burden setting of Haydom, Tanzania.

## OBJECTIVE

The primary objective of this study is to identify potential sources and routes of transmission for *Shigella* and *Campylobacter*, two important causes of diarrhea in young children, in Haydom, Tanzania, with the goal of identifying targeted interventions to confirm the importance of these pathways and reduce exposure to these pathogens.

## METHODS

### Study setting

The study participants will be recruited from four villages surrounding Haydom, Tanzania. The study area has been extensively described^[10,17]^. In brief, Haydom is characterized by poor road infrastructure in a highland setting and primarily dependent on subsistence agriculture and livestock keeping. The area has a high rate of childhood malnutrition and bacterial carriage, previously linked with poor linear growth^[10]^. Understanding how children and the entire household are exposed to these pathogens through the environment is critical for designing targeted and context-specific interventions. The research team has substantial experience with community recruitment, performance of longitudinal cohort studies, sample collection and processing, and the use of molecular methods for detection of enteric pathogens ^[17,18]^.

### Study design and recruitment

This longitudinal cohort study will recruit index children under one year of age as well as their household members, with enrollment occurring over approximately one year and all households followed for 12 months. The index child and all family members who sleep at the house on an average of 4 or more nights per week will be included in the study. Inclusion criteria are 1) the index child must be living in the home, defined by sleeping at the home on average at least 6 nights per week; 2) the family is not planning to move from the study area in next year; and 3) the index child must be below 1 year of age at a time of enrollment and in the current eligible enrollment age stratum (see enrollment below). They will be excluded from the study if 1) the household already has a child enrolled in the study; 2) the index child has been identified to have a developmental abnormality or congenital disease; 3) maternal age is < 18 years; or 4) the index child routinely receives care outside of the home during the day.

### Sample size and power calculation

Assuming that the source analyses are focused on transmission to the index children, and after accounting for the anticipated accrual goal and drop-out rate, a total of 1500 samples from 100 children (13 non-diarrheal and 2 diarrheal stools per index child on average from recent HDSS data) will be included in the analysis. We assumed a design effect of 1.7 to account for correlation between samples from same child, based on the intraclass correlation coefficient of (ICC) 0.05 for *Shigella* and *Campylobacter* detections within samples from the same child in the MAL-ED study^[2]^. We assume the design effect will be similar across *Shigella* and individual *Campylobacter* species. Assuming that 10% of each type of environmental sample tests positive for the target pathogen and the overall prevalence of pathogen infection is 20%, we would have 80% power to detect a risk ratio of 1.76 for each pathogen species and environmental pathway in a two-sided test with alpha=0.05. The minimum detectable risk ratio at 80% power varies by the proportion of environmental samples that test positive for the pathogen and by alternative assumptions for the design effect (Figure 1). Over a range of plausible design effects and pathogen prevalence in varied environmental sample types, we will have power to detect risk ratios that will identify the most important transmission pathways for *Shigella* and the most prevalent individual *Campylobacter* species.

**Figure 1.**
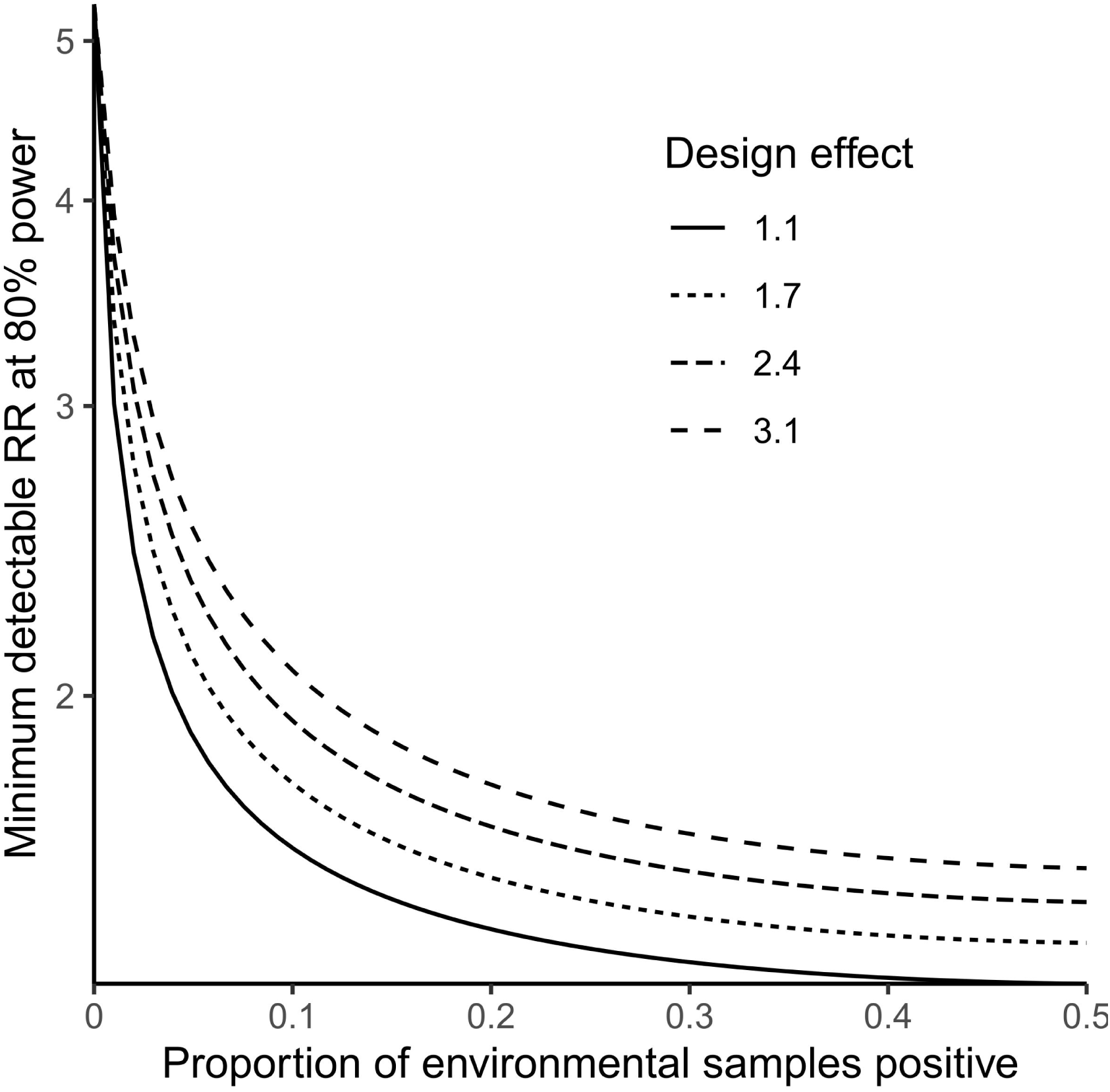
Power calculation for associations between detection of pathogens in the environment and children based on varied assumptions about the design effect and proportion of environmental samples that are positive.

### Enrollment and follow up

To ensure balance of enrolled ages over seasons of the year, enrollment will be stratified by index child age in 3-month strata (0 – 2 months, 3 – 5 months, 6 – 8 months, and 9 – 11 months). Within each 2-week period, one household with a child in each age group will be enrolled. All enrolled index children and their families will be followed for one year from enrollment with twice-weekly visits to identify diarrhea, fever, and antibiotic use (Figure 2). Household stool and environmental samples will be collected monthly, with additional sampling during and after episodes of diarrhea. Diarrhea will be defined by self- or caregiver-report of three or more loose or watery stools in 24 hours or stool with visible blood.

**Figure 2.**
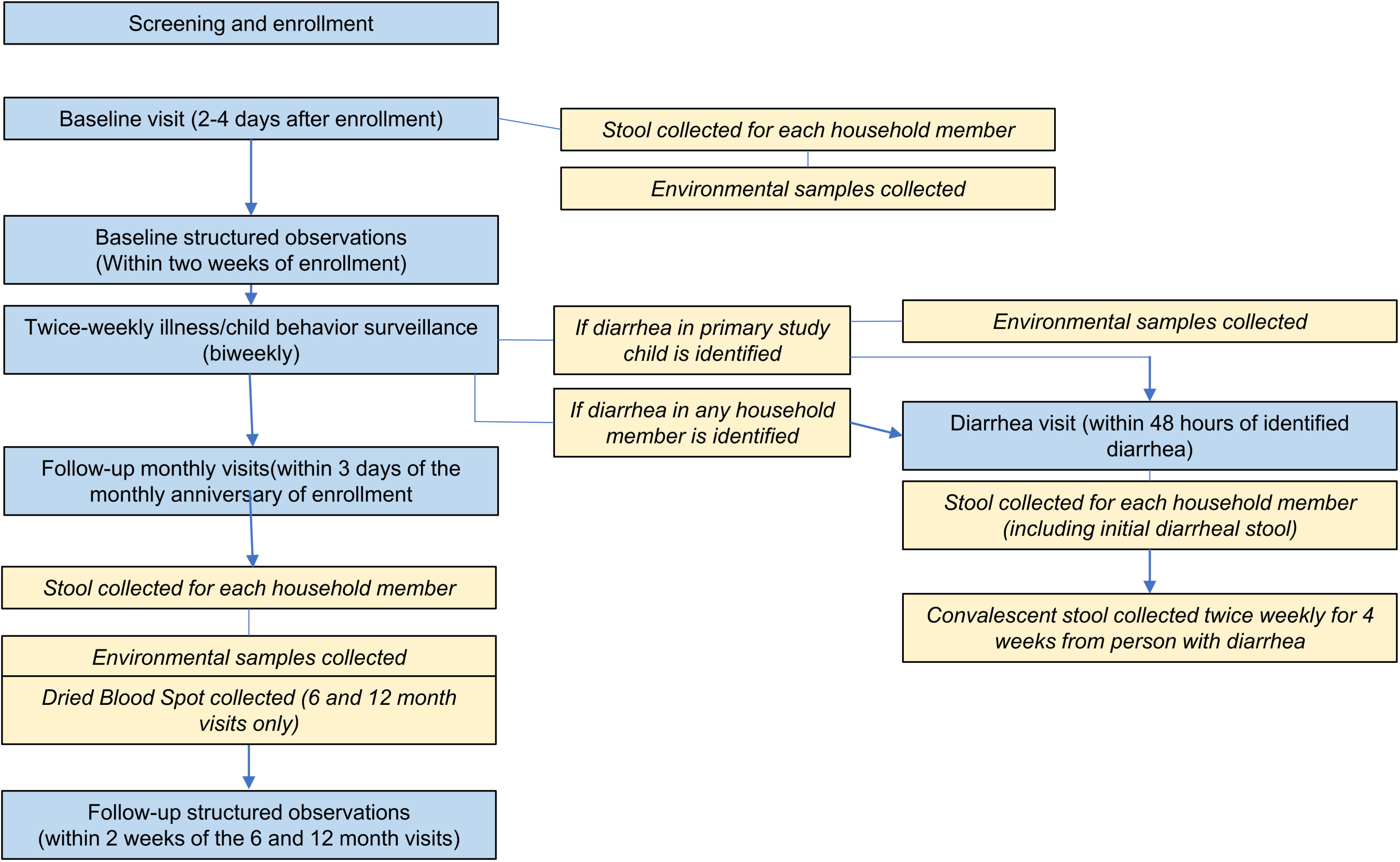
Overview of SCAT study procedures for each enrolled household.

### Characterization of the home environment

At the baseline visit, a semi-structured questionnaire will be used to assess participant sociodemographic characteristics as well as characteristics of the household environment, such as water and sanitation access, house construction materials, presence of handwashing equipment, animal ownership and housing conditions, and food storage and preparation materials at the baseline home visit.

### Structured behavioral observations to quantify pathogen exposures

During the study period, we will conduct direct serial observations of the index children to quantify the potential exposures to *Shigella* and *Campylobacter* through environmental contacts ^[19]^. At baseline, 6 months, and 12 months of follow-up, 6-hour observations on a single day will be conducted by a field staff member who is familiar with the community. The start time of observations will vary from 6 am to noon. Observation will take place only when the child is awake and not defecating in a latrine. During the period of observation, observer will record every contact between a the index child’s mouth and 1) hands ^[8]^, including 2) animals, 3) human and animal feces, 4) water (drinking and swimming/bathing), 5) food, 6) soil, and 7) potential fomites (including cloth and non-cloth objects). We will also document physical contact with other individuals in the household, specifically contact between the child’s mouth and the mother’s breasts or the mother or other individuals’ hands. Behaviors of the primary caregiver will also be recorded, including frequency and duration of conducting activities such as cooking, eating, cleaning, and interacting with children. To record the duration, frequency, and sequence of these contacts, they will be coded using the tablet-based software LiveTrak ^[19]^.

### Monthly household visits and spot checks

During each monthly visit to collect environmental samples, we will conduct a brief household survey and spot-check to record water, sanitation, and hygiene behaviors and indicators of child exposure. Questions include the frequency of observed child ingestion of soil, child mobility, child breastfeeding and feeding practices, animal ownership, animal confinement, animal feces management, frequency and cleaning of animal-raising locations. Spot check observations will include the presence of water storage containers, handwashing stations, soap at handwashing stations, feces on the floor of the latrine, food storage practices, and animals in living spaces, and animal confinement structures.

### Specimen collection, processing, and extraction

We will collect human stool samples as well as multiple environmental specimens from the domestic immediate environment of the index child. Collection, processing and nucleic acid extraction for each sample type is described below. For all samples, Phocine Herpesvirus (PhHV) will be added prior to extraction to assess extraction and amplification efficiency, extraction blanks will be included to monitor extraction contamination, and extracted total nucleic acid (TNA) will be stored at -80°C until PCR is performed.

#### Human stools

Stool samples will be collected upon enrollment, monthly thereafter, and at the time of diarrhea among the index children or any enrolled household member. To characterize duration of pathogen carriage, stool samples from any individual with study-defined diarrhea will continue to be collected approximately twice-weekly for 4 weeks from illness onset, with a collection window of 48 hours. Each participant will collect and provide 5 g or 5 mL of stool sample. All stool samples will be collected within 6 hours of passage and will be transported on ice followed by storage in the lab at -80 °C until TNA extraction. TNA will be extracted using a 200 mg stool aliquot using the QIAamp Fast DNA Stool Mini Kit (Qiagen, Hilden Germany) per manufacturer instructions.

#### Animal feces

At the time of all environmental sampling, fecal samples will be obtained from up to five domestic animal types (cattle, sheep, goats, pigs, dogs and chickens) that are present on the compound of each household. Like in humans, animal stool samples will be transported to the lab on ice and then stored at -80 °C until TNA extraction. Like in human fecal samples, the TNA will be extracted from a 200 mg stool aliquot using a modified the QIAmp Fast DNA Stool mini kit (Qiagen, Hilden Germany) which includes an initial bead beating step followed by the manufacturer’s protocol.

#### Soil

A metal spatula will be used to collect a single soil sample (5 – 10mL) in a 50mL conical tube. This will be collected from all non-cement floors from the kitchen, the sleeping area, the living area, and just outside the primary entrance. Soil samples are not collected if the homes have cement floors. Samples will be refrigerated for up to 18 hours before processing. DNA will be extracted from soil samples using the DNeasy Power Soil Pro Kit (Qiagen, Hilden Germany) per manufacturer instructions.

#### Milk and maize porridge samples

Milk and maize porridge samples will be obtained if they are available in the home on the day of sampling. The caregiver will be asked to serve the milk and porridge or liquid in the way she would give it to the child that day. Up to 5 mL of milk and 10 mL of maize porridge will be collected from the caregiver in a 50mL conical tube. Both milk and maize porridge samples will be placed in a cooler bag with ice packs for transport to the lab. Porridge samples will be homogenized using a bag mixer before DNA extraction. Samples will be refrigerated for up to 18 hours before processing. DNA will be extracted from milk and porridge samples using the QIAamp DNA Mini Kit (Macherey-Nagel, Düren, Germany) per manufacturer instructions.

#### Drinking water

One liter of drinking water will be collected from the household drinking water container and maintained at 4 °C until delivery to the laboratory for concentration and nucleic acid extraction. The water will then be filtered through (up to 3) Argonide filter(s) using a vacuum manifold. The pathogens captured on the filters will then be eluted from the filters using a 1x PBS/0.05 M glycine/ 1% sodium polyphosphate solution, and the eluent will be further concentrated using centrifugal Amicon filters. The final concentrate (approximately 200uL) will be stored at -80°C until TNA extraction, using the QIAmp Fast DNA Stool mini kit (Qiagen, Hilden Germany).

#### Flies

The day prior to each visit for environmental sampling, a fly strip will be distributed to the participants to set up in the home’s cooking area to capture flies over a 24-hour period. At the end of this period, the total number of flies on the tape will be enumerated and field workers will place up to 10 flies into a collection tube and store at -80°C until further processing. The DNA will be extracted from crushed flies using the NucleoSpin® DNA Insect Mini Kit (Macherey-Nagel, Düren, Germany) following manufacturer instructions.

#### Dried Blood Spots

Dried blood spot (DBS) samples will be collected from the index child at home visits by trained study staff via a finger or heel stick to capture 300 µl of whole blood on filter paper in accordance with WHO protocols ^[20]^. DBS will be collected at enrollment and twice during follow-up time points at 6 and 12 months, and within +/- 7 days collection window. DBS will be transferred onto a card, air dried, and kept at room temperature away from direct sunlight until serologic studies are performed ^[21,22]^.

### Detection of *Campylobacter* and *Shigella* by multiplex qPCR

All nucleic acid extracted from human and fecal and from environmental samples will be stored at -80 °C until testing. Testing will consist of a TaqMan-based multiplex qPCR assays to detect *Campylobacter* spp. using species-specific primers and probes targeting the atpA gene^[23]^ and *Shigella* targeting the ipaH gene^[24]^. The assays will be performed on a ViiA7 machine (Thermo Fisher Scientific, Waltham, MA) at an initial heat activation step of 95 °C for 20 min, 40 cycles of two-step PCR including denaturation at 95 °C for 3 seconds, and annealing and extension at 60 °C for 45 seconds.

### Serologic testing

DBS will be tested for IgG against conserved *Shigella* (IpaB) and *Campylobacter jejuni* (p18 and p39) proteins using multiplex bead assays ^[25,26]^. Beads will be stored at 4°C until use, when they will be combined into a multiplex solution and mixed with a concentration of roughly 1000 beads per set per microplate well. DBS will be eluted and added to the solution in each well. The plate will be incubated for 1 hour, after which PE-labeled anti-human IgG will be added and incubated for 1 hour. The plate will be washed with PBS containing 0.05% Tween 20, and then the mean fluorescent intensity will be measured using a Luminex instrument (Luminex Corporation, Austin, Texas, United States).

### Data analysis

The goals of the analysis are to identify possible sources and transmission pathways for *Shigella* and *Campylobacter* using two complementary approaches of statistical and mathematical modeling, whereby the former helps inform the latter. Specifically, we will generate measures of disease incidence, pathway-specific relative risks, environmental contact rates, and duration of pathogen shedding, all of which will inform and parameterize the transmission models. We will then develop complementary transmission models to characterize the relative importance of person-to-person and environmental transmission.

To identify potential sources for infections in index children, we will compare the prevalence of the target pathogen in environmental and familial samples between infected and uninfected index children. We will evaluate possible sources for *Shigella* as well as for *Campylobacter* species identified in at least 5% of stool samples from index children. Source types will be categorized into six pathways with their respective sample types: people (samples: caregiver and sibling stools), animals (animal feces), flies, food, water, and soil. Pathogen burden in each pathway will be characterized as the total qPCR Cq-derived quantity of species-specific pathogen detected in samples from that pathway. The duration of convalescent shedding will be estimated based on the median time from the onset of symptoms to the final positive convalescent stool performed on all qPCR-positive convalescent stool samples.

We will estimate the associations between pathogen quantity in an environmental pathway and infection in the index child using log-binomial regression with generalized estimating equations to account for correlation between samples from the same child over time. We will adjust for age and month of the year given the expected seasonality of exposure. Using the estimated relative risks for each transmission pathway, we will estimate attributable fractions (AF), where AF = 1 – 1/RR and RR is the relative risk for each pathway. We have previously used the attributable fraction methodology to attribute diarrhea etiologies^[18,27]^, a metric that accounts for both the prevalence of detection in a given pathway as well as the strength of association with infection and will provide a rank ordering of the relative importance of each pathway. The estimated relative risks will be used in the transmission models as described below.

The goal of the infection transmission modeling is to understand where the transmission of different enteric pathogens falls on the continuum from primarily person-to-person to primarily environment-to-person transmission, using *Shigella* and *Campylobacter* as model organisms. Our infectious disease transmission modeling approach will consist of two complementary frameworks. First, we will develop an individually based household dynamic transmission model to capture the PCR-confirmed incidence of clinical and subclinical *Shigella* and *Campylobacter* infections observed in this study. Prevalence and duration of shedding in convalescent stools will help to parameterize two model parameters: shedding rate and a factor converting shedding from humans into increases in environmental contamination. Second, we will use a compartmental model framework to develop an endemic transmission model of the cohort. We will incorporate the prevalence of collected household characteristics, environmental contacts and measured environmental exposures (water, food, flies, etc.). Finally, relative risks estimated from the associations between detections in environmental and familial samples and infections in the index child will provide informative priors for parameters that characterize the importance of person-to-person and environmental transmission.

## Data Availability

Data will be made available in a public repository.

## ETHICS AND DISSEMINATION

The study was granted an ethical approval and registered by the National Institute for Medical Research (NIMR) in Tanzania (NIMR/HQ/R.8a/Vol.IX/4023) and the University of Virginia Institutional Review Board (HSR210431) in the US. Permission to conduct the study was granted by the local leaders in the study catchment area and the HLH administrative authorities. Prior to any study procedure, all members of eligible households will consent to participate in the study, provide sociodemographic, clinical information, specimens for laboratory testing and their microbiological information to be used in the study. Each household approached for participation will be informed about the study, and if found eligible for enrollment will go through a voluntary consent process. This will include informed consent of adults and of parents/guardians for children under 18 years of age. For children 7-17 years of age, verbal assent will also be obtained. For both the consent and assent, participants will have the opportunity to ask questions and review the study information and consent/assent forms. They do not need to consent/assent right away but will have time to think about their response and ask any questions about the protocol/study. They will be encouraged to seek advice from friends and family before signing consent. All subjects will be asked open-ended questions to confirm that they understand the study. For the potential participants who are illiterate, they will be allowed to identify an impartial witness who will participate in the consenting process. If they understand and consent to participation in the study, they will mark with a thumbprint and the impartial witness will sign the consent. A copy of the signed informed consent form will be given to all participants.

The scientific reports resulting from this study will be made publicly available, through publication in open-access journals and conference presentations. Data will be made available in a public repository. An integrative knowledge translation strategy will be used for involving policy makers and key stakeholders at the Ministry of Health in Tanzania and beyond.

## Author’s contributions

JNSE, ETRM, MT, EM, and JAPM conceived of the study and obtained funding. PMM and JAPM drafted the manuscript. AFB, JNSE, ETRM developed the analysis plan with imput from SE and GWG. EGW, SP, TA, MT, FH, CK, KM, and RM developed and refined the sampling, processing, extraction, and testing protocols. SK, SE, GWG, ETRM, EM, and JAPM developed the study protocol, procedures, and data collection instruments. All authors contributed to the development of this manuscript and/or study procedures and to reading and approving the final version for publication.

